# Evaluation of a commercial 16S rDNA PCR assay (UMD-SelectNA, Molzym) for pathogen detection from cardiac tissue specimens

**DOI:** 10.1101/2025.04.03.25325084

**Authors:** Manuel Wolters, Konstantin Tanida, Flaminia Olearo, Benjamin Berinson, Christoph Sinning, Martin Aepfelbacher, Holger Rohde, Martin Christner

**Author notes:** authors contributed equally to work (Manuel Wolters, Konstantin Tanida).

## Abstract

The detection of bacteria in cardiac tissue, vegetations, or explanted prosthetic valves using nucleic acid amplification tests, such as 16S rDNA amplicon sequencing (16S rDNA PCR), has been incorporated as pathologic criterion for definite infective endocarditis into the 2023 Duke-International Society for Cardiovascular Infectious Diseases Criteria for Infective Endocarditis. To verify results from previous studies suggesting increased detection rates and to investigate the added diagnostic value of 16S rDNA PCR, we prospectively compared the diagnostic yield of a CE-IVD-marked 16S rDNA PCR assay to conventional culture (CC) in a large number of cardiac tissue samples, relating our findings to microbiological evidence from blood culture sampling and patient history. Over a seven-year period (2016-2023), 687 cardiac tissue samples were subjected to CC and 16S rDNA PCR. 16S rDNA PCR and CC yielded positive results in 326 (47.4 %) and 154 (22.4 %) samples, respectively. 136 (19.7 %) samples were concordantly tested positive. In 190 (27.7 %) and 18 (2.6 %) samples, a positive result was obtained only by PCR or CC. 343 (49.9 %) samples tested negative by both methods. In a case-based analysis of a subgroup of blood culture-confirmed IE (241 cases) and BC-negative non-IE cases (126 cases), 16S rDNA PCR yielded a diagnostic sensitivity and specificity of 74.7 % and 98.4 %, while CC yielded a sensitivity and specificity of 38.6 % and 99.2 %. In conclusion, the use of 16S rDNA PCR significantly increased the detection rate for bacteria in heart valve tissue. A notable proportion of the culture-negative samples contained pathogens that are generally not detectable by conventional culture. Therefore, supplementation of CC with 16S rDNA PCR is recommended for cardiac tissue samples, especially in case of blood culture-negative endocarditis.

## Introduction

Infective endocarditis (IE) is a life-threatening disease that, without prompt and appropriate treatment, can result in serious heart damage or death ^1–3^. Even with adequate treatment, more than a third of patients dies within a year of diagnosis ^4^. Establishing the causative microorganism and its susceptibility profile plays a crucial role in confirming the diagnosis and optimizing antibiotic treatment. Blood cultures are the standard test for determining the microbiological etiology of IE, as they provide live bacteria for both identification and susceptibility testing ^1,2,5^. However, in around 10 to 20 % of cases, blood cultures remain negative, a condition referred to as blood culture-negative infective endocarditis (BCNIE) ^6–8^. BCNIE can occur due to previous antibiotic administration or the presence of fastidious bacteria that grow poorly (e.g., *Cutibacterium spp*.) or do not grow at all in conventional blood culture media (e.g., *Bartonella spp*., *Tropheryma whipplei, Coxiella burnetii*). In some cases of BCNIE, the causative organism can be identified by serologic testing (*C. burnetii, Bartonella spp*.) or by pathogen-specific PCR from blood (*C. burnetii*) ^5^.

Surgical intervention is required in 25 % to 53 % of cases of endocarditis ^3^, and excised cardiac tissue is a valuable specimen for microbiological evaluation, particularly in cases of BCNIE. However, cardiac tissue culture, has a relatively low sensitivity, with positivity rates ranging from only 12 % to 40 % ^9–17^. In recent years, nucleic acid detection from excised heart tissue has emerged as a promising technique for pathogen detection and IE diagnosis. PCR amplification and sequencing of the 16S rRNA gene (16S rDNA PCR) is an agnostic approach that allows for the detection of all potential bacterial pathogens, including rare, fastidious and non-cultivable agents, while not being limited to a certain set of target organisms like syndromic PCR panels. Several studies have demonstrated superior clinical sensitivity of 16S PCR compared to valve culture, but sample sizes were often limited, and a variety of DNA extraction and amplification protocols were used, limiting comparability ^9,11–16,18–23^. Nonetheless, the identification of microorganisms from cardiac tissue, vegetations, or explanted prosthetic valves by 16S PCR was recently included as pathologic criterion for definite IE in the 2023 Duke-International Society for Cardiovascular Infectious Diseases Criteria for IE ^24^.

In this study, we report the evaluation of the only currently available European certificate of conformity (CE) marked 16S rDNA PCR assay (SepsiTest, UMD-SelectNA, Molzym, Germany) for pathogen detection from tissue samples in a large series of cardiac tissue specimens from IE and non-IE patients.

## Methods

### Study setting

This study was conducted to compare the performance of bacterial pathogen detection by 16S rDNA PCR with conventional culture (CC) from cardiac tissue samples sent to our microbiology laboratory at the University Medical Center Hamburg-Eppendorf, a tertiary care center with 1600 beds. From 2016 to 2023, 687 samples from 570 cases (range: 1-4 samples per case) were processed simultaneously using both methods as part of routine diagnostics (Figure 1). For the case-based analysis, a case was defined as a hospitalization in which at least one cardiac tissue sample was obtained and sent in for pathogen detection. A case was considered positive by 16S rDNA PCR or CC when at least one cardiac sample tested positive using the respective technique. A retrospective chart review of these cases was conducted to obtain information on blood culture results and the clinical pre-surgery classification as IE or non-IE cases, as determined by the treating physicians. When no clear information on the diagnosis (IE/non-IE) was available, the case was classified as unresolved.

**Figure 1:**
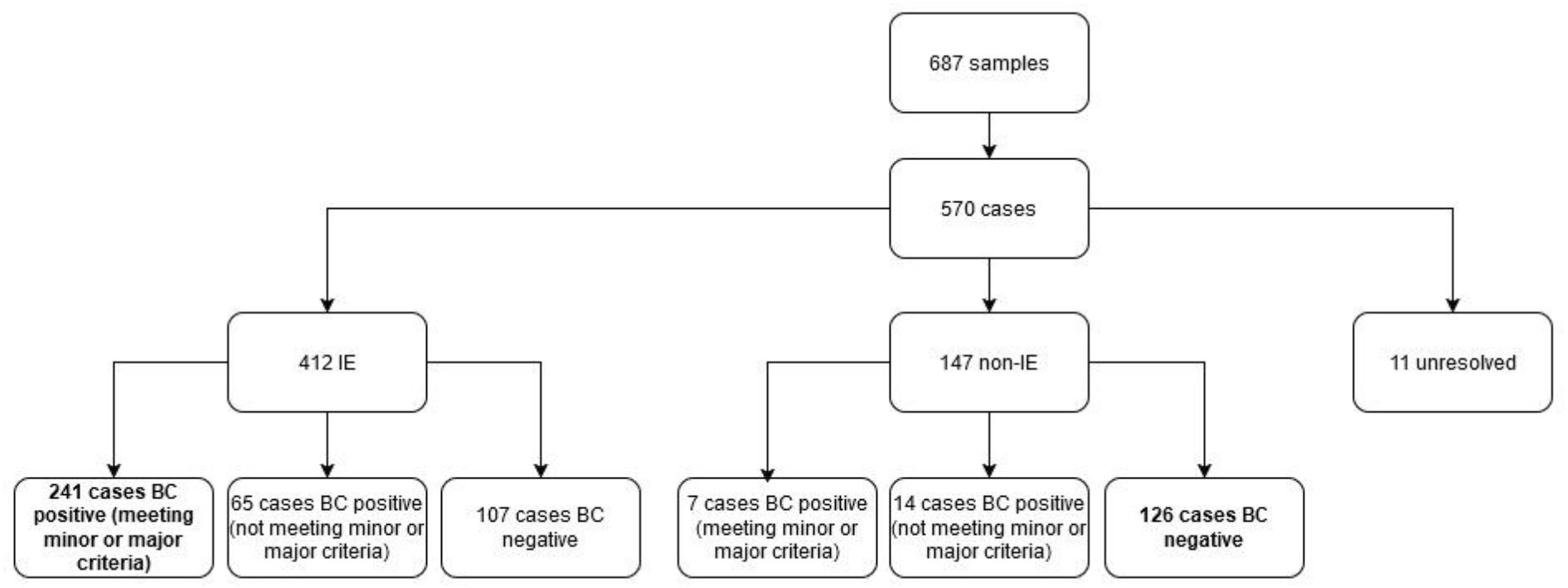
Flowchart of included cardiac tissue samples and respective cases, categorized by clinical diagnosis of IE. Bold letters indicate the subgroups used for diagnostic performance analysis (Table 3).

### Sample processing

Native, biological and mechanical heart valves, vegetations and abscess material was homogenized in Ultra-Turrax mixer tubes (Ultra-Turrax Tube-Drive; IKA Werke GmbH & Co. KG, Staufen, Germany) with sterile saline.

### Conventional culture

Homogenized samples were inoculated onto Columbia sheep blood agar, Chocolate agar, Schaedler agar and into thiogylocolate broth (all Oxoid; Basingstoke, UK). Columbia and chocolate agar plates were incubated at 37 °C in 5 % CO_2_ for 14 days, with daily inspection until day 3, followed by additional readings after 7, 10, and 14 days of incubation. Schaedler agar plates were incubated at 37 °C under anaerobic conditions and inspected on day 3. Broth cultures were incubated at 37 °C in ambient air, and inspected daily for 3 days. Broths were streaked only in case of visible growth. Bacterial isolates were identified to the species level using MALDI-TOF mass spectrometry fingerprinting with a Biotyper instrument according to the manufacturer’s instructions (Maldi Biotyper; Bruker, Bremen). Additional 16S rDNA sequencing was performed as required.

### 16S rDNA PCR

One milliliter of homogenized tissue was subjected to human DNA depletion and bacterial DNA extraction using the SelectNA plus robotic system run with the MolYsis-SelectNA plus kit (Molzym, Bremen, Germany) according to the manufacturer’s instructions. In brief, human and dead bacterial cells were lysed under chaotropic conditions and the released DNA was degraded by DNase. Subsequently, intact microorganisms were collected and subjected to microbial DNA extraction. SYBR-Green-based 16S rDNA PCR with Mastermix 16S complete (Molzym; Bremen, Germany) was performed on a LightCycler480 instrument (Roche, Basel, Switzerland). Amplification and melt curves were analyzed manually. Samples with successful PCR amplification (Cq <= 35) and a melting temperature between 86 °C and 92 °C were considered positive. PCR eluates from positive samples were purified (QIAquick PCR purification kit; Qiagen, Hilden, Germany) and sent for Sanger sequencing. 16S sequences were analyzed with the web-based SepsiTest-BLAST application provided by the assay manufacturer. If the SepsiTest application provided no identification, the respective sequences were additionally tested against NCBI’s 16S database.

## Results

### Sample-based analysis

Results from 16S rDNA PCR and CC of 687 samples are summarized in Table 1 (complete dataset in Table S1). 16S rDNA PCR and CC yielded positive results in 326 of 687 (47.4 %) and 154 of 687 (22.4 %) cardiac tissue samples, respectively. The detected species are summarized in Table 2 and correspond to the expected pathogen spectrum in cases of IE for both methods. 136 of 687 (19.7 %) samples tested positive with both methods. In 134 of these samples, concordant species identification was obtained. The two discordant samples yielded likely contaminants by CC (sparse growth of CoNS on solid medium and growth of *Enterococcus faecium* in enrichment broth only) while 16S rDNA PCR identified typical IE pathogens (*Staphylococcus aureus* and *Streptococcus salivarius* group). In 190 of 687 (27.7 %) and 18 of 687 (2.6 %) samples positive results were exclusively obtained by 16S rDNA PCR or CC (Table 2). 343 of 687 (50.0 %) samples tested negative with 16S rDNA PCR and CC.

**Table 1:** Results of 16S rDNA PCR and CC in 687 cardiac tissue samples.

|  | CC negative | CC positive | Total |
| --- | --- | --- | --- |
| **16S rDNA PCR negative** | 343 (49.9 %) | 18 (2.6 %) | 361 (52.5 %) |
| **16S rDNA PCR positive** | 190 (27.7 %) | 136 (19.8 %) | 326 (47.5 %) |
| **Total** | 533 (77.6 %) | 154 (22.4 %) | 687 (100.0 %) |

**Table 2:**
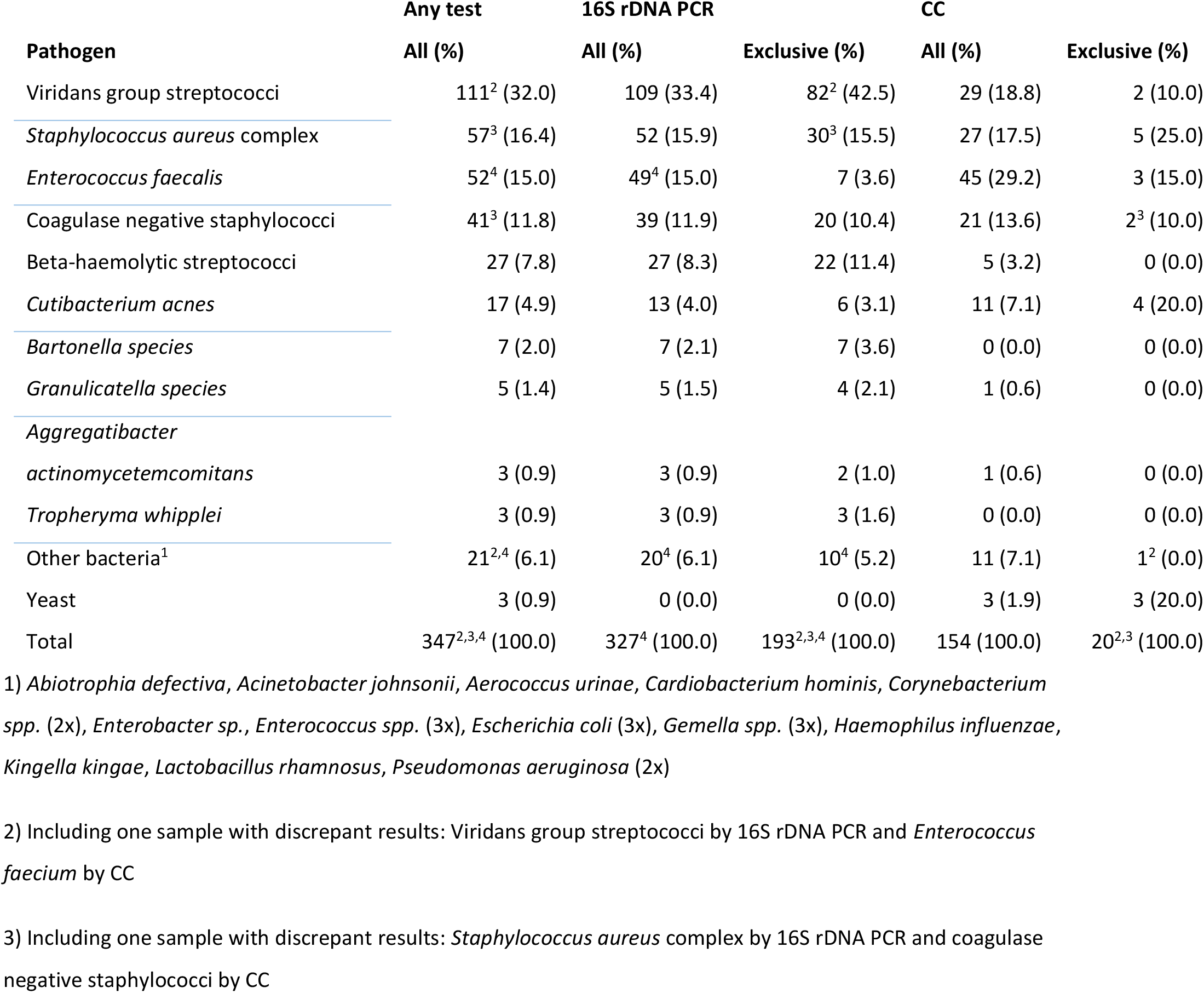
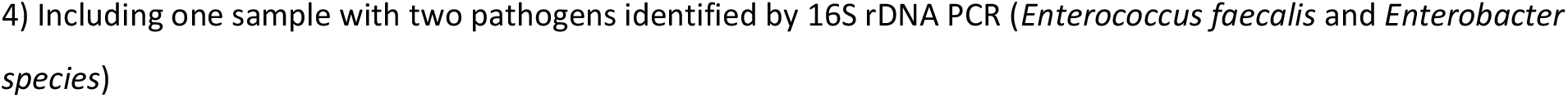
Pathogens identified by 16S rDNA PCR or CC in 344 positive cardiac tissue samples.

| Pathogen | Any test | 16S rDNA PCR |  | CC |  |
| --- | --- | --- | --- | --- | --- |
|  | All (%) | All (%) | Exclusive (%) | All (%) | Exclusive (%) |
| Viridans group streptococci | 111<sup>2</sup> (32.0) | 109 (33.4) | 82<sup>2</sup> (42.5) | 29 (18.8) | 2 (10.0) |
| *Staphylococcus aureus* complex | 57<sup>3</sup> (16.4) | 52 (15.9) | 30<sup>3</sup> (15.5) | 27 (17.5) | 5 (25.0) |
| *Enterococcus faecalis* | 52<sup>4</sup> (15.0) | 49<sup>4</sup> (15.0) | 7 (3.6) | 45 (29.2) | 3 (15.0) |
| Coagulase negative staphylococci | 41<sup>3</sup> (11.8) | 39 (11.9) | 20 (10.4) | 21 (13.6) | 2<sup>3</sup> (10.0) |
| Beta-haemolytic streptococci | 27 (7.8) | 27 (8.3) | 22 (11.4) | 5 (3.2) | 0 (0.0) |
| *Cutibacterium acnes* | 17 (4.9) | 13 (4.0) | 6 (3.1) | 11 (7.1) | 4 (20.0) |
| *Bartonella species* | 7 (2.0) | 7 (2.1) | 7 (3.6) | 0 (0.0) | 0 (0.0) |
| *Granulicatella species* | 5 (1.4) | 5 (1.5) | 4 (2.1) | 1 (0.6) | 0 (0.0) |
| *Aggregatibacter* |  |  |  |  |  |
| *actinomycetemcomitans* | 3 (0.9) | 3 (0.9) | 2 (1.0) | 1 (0.6) | 0 (0.0) |
| *Tropheryma whippelii* | 3 (0.9) | 3 (0.9) | 3 (1.6) | 0 (0.0) | 0 (0.0) |
| Other bacteria<sup>1</sup> | 21<sup>2,4</sup> (6.1) | 20<sup>4</sup> (6.1) | 10<sup>4</sup> (5.2) | 11 (7.1) | 1<sup>2</sup> (0.0) |
| Yeast | 3 (0.9) | 0 (0.0) | 0 (0.0) | 3 (1.9) | 3 (20.0) |
| **Total** | **347<sup>2,3,4</sup> (100.0)** | **327<sup>4</sup> (100.0)** | **193<sup>2,3,4</sup> (100.0)** | **154 (100.0)** | **20<sup>2,3</sup> (100.0)** |

1) *Abiotrophia defectiva*, *Acinetobacter johnsonii*, *Aerococcus urinae*, *Cardiobacterium hominis*, *Corynebacterium* spp. (2x), *Enterobacter* sp., *Enterococcus* spp. (3x), *Escherichia coli* (3x), *Gemella* spp. (3x), *Haemophilus influenzae*, *Kingella kingae*, *Lactobacillus rhamnosus*, *Pseudomonas aeruginosa* (2x)
2) Including one sample with discrepant results: Viridans group streptococci by 16S rDNA PCR and *Enterococcus faecium* by CC
3) Including one sample with discrepant results: *Staphylococcus aureus* complex by 16S rDNA PCR and coagulase negative staphylococci by CC
4) Including one sample with two pathogens identified by 16S rDNA PCR (*Enterococcus faecalis* and *Enterobacter species*)

### Case-based analysis

To evaluate the clinical performance of 16S rDNA PCR and CC, blood culture results as well as clinical information were retrospectively collected by patient chart review (570 cases) (Figure 1). In 247 of 570 cases (43.3 %), positive BC results were documented that met the Duke major or minor criteria. Results from blood culture were concordant in 98.9 % of cases with positive 16S rDNA PCR (180 cases) and 96.8 % of cases with positive CC (93).

A clinical diagnosis of infective endocarditis was documented in 412 of 570 cases (72.3 %) and in 147 of 570 (25.8 %) an alternative diagnosis (mostly valvular heart disease due to non-infectious conditions, e.g., degenerative) was found, while 11 cases were classified as unresolved. To obtain an estimate of clinical sensitivity and specificity of 16S rDNA PCR and CC, a subgroup of blood-culture positive IE cases (clinical diagnosis of IE and BC results meeting Duke major or minor criteria, 241 cases) and BC-negative non-IE cases (126 cases) were analyzed. In this subgroup 16S rDNA PCR yielded a sensitivity and specificity of 74.7 %, and 98.4 %, respectively, as compared to 38.6 % and 99.2 % obtained with CC (Table 3). Among the 126 cases that were clinically categorized as non-IE, only two cases had a positive result in the 16S rDNA PCR detecting *Streptococcus mitis* group and *Streptococcus dysgalactiae*. In the first case, a *Streptococcus mitis* endocarditis seven months prior to cardiac surgery was documented and in the second case, intraoperative valve inspection was also suggestive for a previous IE.

**Table 3:** Diagnostic performance of 16S rDNA PCR and CC in a case-based analysis of a subgroup of 241 blood culture confirmed IE and 126 BC-negative non-IE cases.

|  | PCR |  | CC |  |
| --- | --- | --- | --- | --- |
|  | Value | 95% CI | Value | 95% CI |
| Sensitivity | 74.7 % | 68.7 % to 80.1 % | 38.6 % | 32.4 % to 45.1 % |
| Specificity | 98.4 % | 94.4 % to 99.8 % | 99.2 % | 95.7 % to 100 % |
| Disease prevalence | 65.7 % | 60.6 % to 70.5 % | 65.7 % | 60.6 % to 70.5 % |
| Positive Predictive Value | 98.9 % | 95.8 % to 99.7 % | 98.9 % | 92.9 % to 99.9 % |
| Negative Predictive Value | 67.0 % | 62.0 % to 71.7 % | 45.8 % | 43.3 % to 48.3 % |
| Accuracy | 82.8 % | 78.6 % to 86.6 % | 59.4 % | 54.2 % to 64.5 % |

Importantly, in 41 of 412 IE cases (10.0 %), 16S rDNA PCR was the only test yielding a presumptive pathogen identification; this number increased to 69 cases (16.5 %) if BC was required to meet the Duke major or minor criteria. In 9 cases, bacterial species that could not have been detected by BC or CC (n=6 *Bartonella species*, n=3 *Tropheryma whipplei*) were identified. The distribution of pathogens is shown in Table 4.

**Table 4:** Pathogens exclusively identified by 16S rDNA PCR in 412 cases of IE.

| Pathogen | BC negative or not meeting Duke microbiologic criteria (% of all cases) |  | BC negative (% of all cases) |
| --- | --- | --- | --- |
| Viridans group streptococci | 25 (6.1) |  | 16 (3.9) |
| Beta-haemolytic streptococci | 10 (2.4) |  | 6 (1.5) |
| Coagulase-negative staphylococci | 9 (2.2) |  | 3 (0.7) |
| *Bartonella species* | 5 (1.2) |  | 5 (1.2) |
| *Cutibacterium acnes* | 4 (1.0) |  | 4 (1.0) |
| *Enterococcus faecalis* | 4 (1.0) |  | 2 (0.5) |
| *Tropheryma whipplei* | 3 (0.7) |  | 3 (0.7) |
| *Staphylococcus aureus complex* | 2 (0.5) |  | 0 (0.0) |
| *Acinetobacter johnsonii* | 1 (0.2) |  | 1 (0.2) |
| *Cardiobacterium hominis* | 1 (0.2) |  | 0 (0.0) |
| *Corynebacterium species* | 1 (0.2) |  | 0 (0.0) |
| *Enterococcus species* | 1 (0.2) |  | 0 (0.0) |
| *Granulicatella adiacens* | 1 (0.2) |  | 0 (0.0) |
| *Haemophilus parainfluenzae* | 1 (0.2) |  | 1 (0.2) |
| *Kingella kingae* | 1 (0.2) |  | 0 (0.0) |
| **Total** | **69 (16.7)** |  | **41 (10.0)** |

## Discussion

Identification of the causative pathogen is pivotal for optimized treatment of IE patients but fails in a substantial number of cases when BC remains negative due to previous antibiotic treatment or in the presence of difficult-to-culture species. In cases where valve surgery is required, examination of excised cardiac tissue offers an additional chance to identify the etiologic agent of IE. This study investigated the performance of a CE-IVD-marked 16S rDNA PCR assay compared to CC for IE pathogen detection from cardiac samples in a large single-center cohort.

Our data demonstrate that positive 16S rDNA PCR results are highly accurate in detecting the true causative agent of IE, as the identified bacterial species were highly concordant with identifications from CC (98.5 %) and BC (98.9 %).

We also found a more than twofold higher positivity rate of 16S rDNA PCR (47.4 %) as compared to CC (22.4 %), confirming previous evidence that 16S rDNA PCR can substantially increase pathogen detection rates from cardiac tissue samples. In addition, the distribution of detection rates for 16S rDNA PCR and CC implied species-specific performance differences between both techniques. As expected, PCR was superior to CC for fastidious organisms like *Bartonella, Granulicatella, Aggregatibacter* and *Tropheryma whipplei*. A similar pattern, albeit to a lesser extent, was also observed for the non-fastidious beta-hemolytic and Viridans group streptococci, probably due to the detrimental effect of antimicrobial treatment on CC yield. In accordance with an assumed treatment effect, comparatively higher CC detection rates were found for *Staphylococcus aureus*, coagulase negative staphylococci, *Enterococcus faecalis* and *Cutibacterium acnes*; species which had been associated with treatment failure and increased relapse rates in IE ^25,26^. Detection rate profiles for *Staphylococcus aureus* and *Cutibacterium acnes* were also compatible with reduced PCR detection rates.

On the other hand, patient chart review revealed that a relevant proportion of cases with clinically diagnosed blood culture positive IE had negative 16S rDNA PCR results (61 of 241, 25.3 %) suggesting suboptimal diagnostic sensitivity of the employed 16S rDNA PCR assay. Although a systematic analysis with tissue samples had not been performed, the test is unlikely to outperform CC in terms of analytical sensitivity, given the limits of detection reported for whole blood specimen ^27^. Accordingly, several studies have associated false negative results with low bacterial counts in the corresponding cultures ^28,29.^

Molecular assays which might overcome the perceived limitations of 16S rDNA PCR include species-specific qPCR panels and next-generation sequencing (NGS) approaches. While qPCR is promising in terms of analytical sensitivity and turnaround time, suitable assays have not yet been specifically developed or evaluated for the use with cardiac tissue. Apart from analysis of valve tissue, shotgun metagenomic sequencing of microbial cell-free DNA (mcfDNA) and NGS-based analysis of 16S rDNA PCR amplicons have recently been proposed as non-invasive tests for IE ^30–32^. In a small cohort, both techniques showed similar detection rates, allowing pathogen identification in cases of BCNIE ^31^. However, larger prospective studies are needed to establish clinical performance, added value and specific indication for these expensive tests. Despite the promising features offered by novel molecular assays, it should not be forgotten that a low false positivity rate is of utmost importance for tests performed to inform targeted antimicrobial treatment. Consequently, careful balancing sensitivity and specificity will be critical as molecular testing becomes more widely used in the clinical diagnosis of IE.

The present study has several limitations, such as the monocentric approach and the retrospective collection of clinical data by patient chart review. Due to partially incomplete documentation, it was not possible to strictly classify patients according to the Duke criteria, but IE classification was based on the judgement of the treating physicians and blood culture results as documented in the patient records. In addition, 16s rDNA PCR and CC were performed as part of routine diagnostics, so that the results from both assays were not blinded to laboratory staff.

## Conclusion

16s rDNA PCR is highly accurate for the detection of the causative agent of IE in explanted cardiac tissue, offering higher positivity rates and diagnostic sensitivity than CC while retaining excellent specificity. Tissue cultures should thus regularly be supplemented with 16s rDNA PCR, especially in cases of BCNIE.

## Data Availability

All data produced in the present study are available upon reasonable request to the authors.

## Acknowledgements

The authors thank the staff at the Institute for Medical Microbiology, Virology and Hygiene for excellent technical assistance.

## Statements and Declarations

The authors have no competing interests to declare

## Notes

### Competing Interest Statement

The authors have declared no competing interest.

### Funding Statement

This study did not receive any funding.

### Author Declarations

According to the Ethics Committee of the Hamburg Chamber of Physicians, no informed consent was required for the collection, analysis, and publication of these data.

